# Labeling and Disclosure of AI-generated Mental Health Content on TikTok

**DOI:** 10.64898/2026.08.24.26361037

**Authors:** Alex Christiansen, Ruth Page

## Abstract

TikTok has become a significant source of health information, and concern has grown about AI-generated content (henceforth, ‘AIGC’) as a vehicle for health misinformation. Where AIGC presents realistic-appearing people giving health advice, disclosure labels are the viewer’s only reliable cue that what they are watching is synthetic. This research letter compares AI label metadata across 128,016 mental health-related TikTok videos and 4,924 videos from a network of 50 profiles posting exclusively AI-generated mental health content to evaluate how much content reaches audiences undisclosed. In a keywords-based collection, fewer than a percent of TikTok videos about mental health carried an AI label, but in profiles containing purely AI-generated content, just over 9 in 10 videos (90.23%) were neither labelled by the creator nor identified by TikTok’s automatic detection. Additionally, in the keyword collection, automatic detection produced the majority of labels, while in confirmed AI-generated content from 50 profiles, it accounted for just three of the 481 labelled videos. These findings highlight the challenging landscape of AI disclosure and labelling and raise questions about where automatic detection is failing.

## Introduction

TikTok has become a significant source of health information, and concern has grown about AI-generated content (henceforth, ‘AIGC’) as a vehicle for health misinformation ^1^. Where AIGC presents realistic-appearing people giving health advice, disclosure labels are the viewer’s only reliable cue that what they are watching is synthetic. Both recent EU regulations and TikTok’s policies require creators to label “realistic-appearing scenes or people” and prohibit “fake authoritative sources” ^2^. TikTok reported 1.3 billion pieces of content tagged as AIGC in November 2025, rising to 3 billion by July 2026 ^3,4^, a small share for a platform that averaged over 100 million daily uploads in 2025 ^5^. To evaluate how much AI-generated content reaches audiences undisclosed, we analyzed AI label metadata in two datasets of TikTok videos covering mental health.

## Methods

The TikTok Research API contains AI labels as metadata, shown as either creator-labelled, where the creator has tagged the content as consisting of or containing AI-generated elements, or automated, where TikTok’s systems have identified and applied the label. Both are limited to true positives and provide no view of AI incorrectly remaining unlabeled.

### Keywords

To estimate AI labelled mental health content, we collected and analyzed metadata from more than 128,000 TikTok videos in the eight-month period 09/2025-04/2026 wherein the ‘video description’ text mentioned at least one of five common mental health conditions (adhd; anxiety; bipolar; depression; ptsd) or a popular general mental health hashtag (mentalhealth; mentalhealthawareness; mensmentalhealth).

### Profiles

To estimate unlabeled AIGC, we used labelled content from the keywords collection to identify a network of 50 profiles containing only realistic, AI generated videos. Candidates were examined manually for converging indicators of centralized, templated production, by identifying (a) identical or near-identical branding/logos across profile imagery, (b) verbatim reuse of verbal audio scripts across profiles, (c) repost of identical video files across ostensibly distinct profiles. We collected metadata from all 4,924 videos produced by the profiles in two waves, in December 2025 and April 2026. Content centered on mental health, but frequently overlapped mentions of anxiety, depression, stress and disordered eating with gut health, hormonal imbalance and liver damage as part of promotions of supplements.

## Results

As shown in Table 1, 0.86% of videos (n = 1,095) in the keywords collection carried an AIGC label. Of those, 0.39% (n = 496) were applied by creators and 0.47% (n = 599) by automatic detection. In the 50 AIGC-exclusive profiles, only 9.77% (n = 481) of 4,924 videos were correctly labelled, leaving over nine in ten (90.23%, n = 4,443) of confirmed AI-generated videos undisclosed. Automatic detection accounted for just 3 of the 481 labels (0.06%).

**Table 1.**
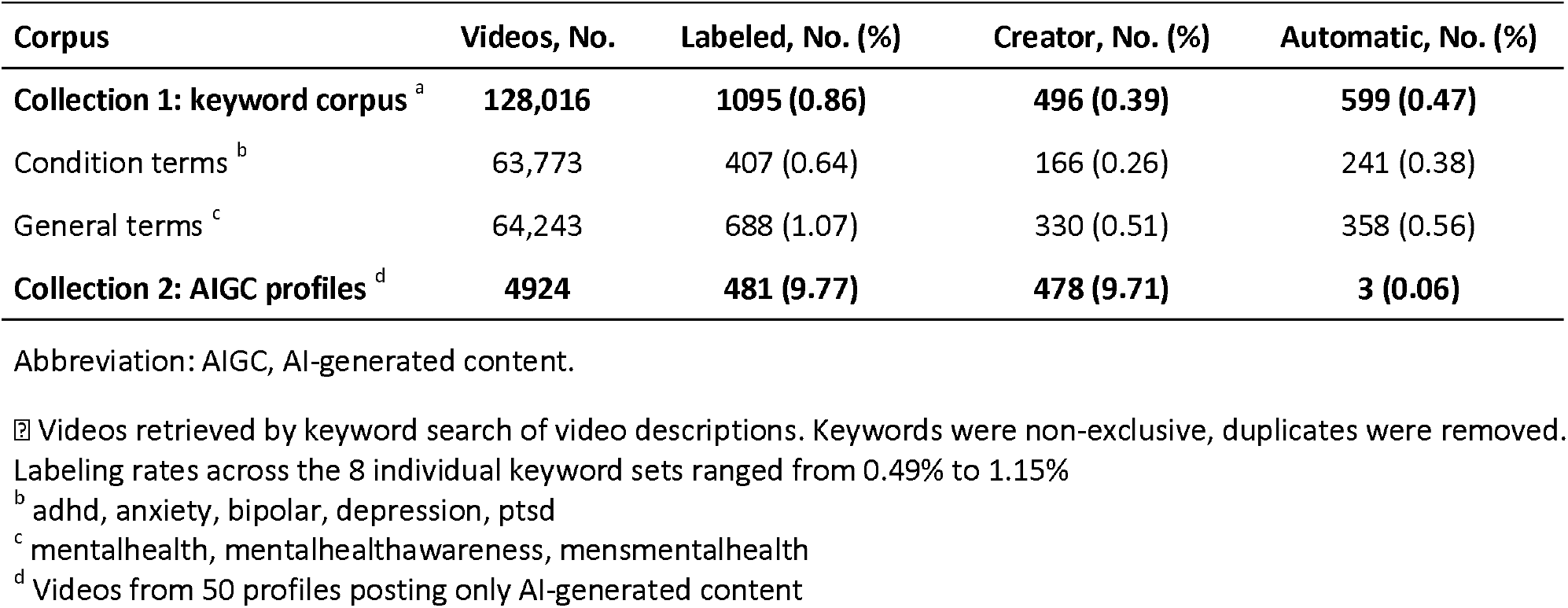
Creator-Applied and Automatic AI Labeling in Two Collections of Mental health Content From TikTok.

| Corpus | Videos, No. | Labeled, No. (%) | Creator, No. (%) | Automatic, No. (%) |
| --- | --- | --- | --- | --- |
| **Collection 1: keyword corpus <sup>a</sup>** | **128,016** | **1095 (0.86)** | **496 (0.39)** | **599 (0.47)** |
| Condition terms <sup>b</sup> | 63,773 | 407 (0.64) | 166 (0.26) | 241 (0.38) |
| General terms <sup>c</sup> | 64,243 | 688 (1.07) | 330 (0.51) | 358 (0.56) |
| **Collection 2: AIGC profiles <sup>d</sup>** | **4924** | **481 (9.77)** | **478 (9.71)** | **3 (0.06)** |

Abbreviation: AIGC, AI-generated content.
<sup>a</sup> Videos retrieved by keyword search of video descriptions. Keywords were non-exclusive, duplicates were removed. Labeling rates across the 8 individual keyword sets ranged from 0.49% to 1.15%
<sup>b</sup> adhd, anxiety, bipolar, depression, ptsd
<sup>c</sup> mentalhealth, mentalhealthawareness, mensmentalhealth
<sup>d</sup> Videos from 50 profiles posting only AI-generated content

Labelling strategies differed significantly between profiles. Nearly half (48%, n = 24) labelled none of their content, while ten profiles accounted for 83.4% of all labels. As shown in Figure 1, video production within the network continued across the full 2025-2026 period, but labelling did not. This effect was consistent across profiles, whether they had labelled heavily or minimally prior to this point. Among the 29 profiles that were active in both 2025 and 2026, labelling fell from 12.50% to 2.67%.

**Figure 1.**
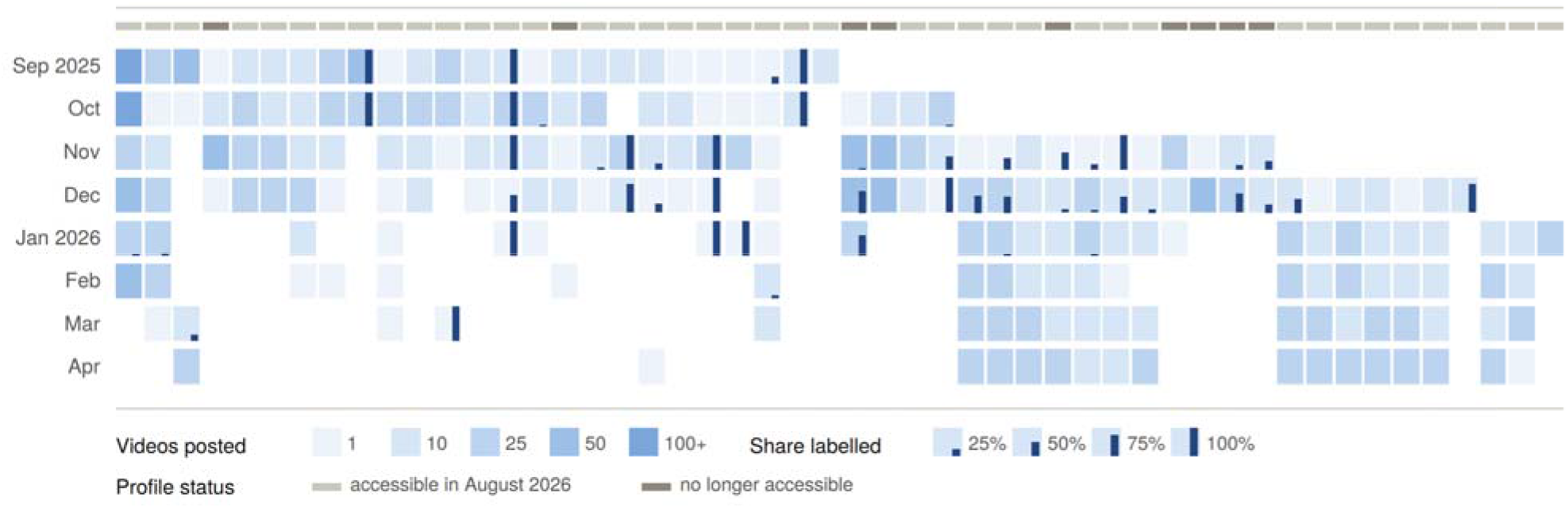
Monthly Video Production and AI Labelling Across 50 Networked AIGC Profiles. Heatmap showing posting frequency and AIGC labelling consistency across 50 profiles in an 8-month span. Cell shading indicates quantity of videos posted; empty cells suggest no posts. The bar within each cell indicates the proportion of posts carrying an AI label.

## Discussion

Our primary finding is that AI labels appear to be an unreliable indicator of synthetic content for viewers seeking mental health information on TikTok. Fewer than 1% of mental health videos retrieved by keyword carried a label, yet in a collection made up entirely of AIGC, over nine in ten videos were neither disclosed by creators nor detected by TikTok’s automated systems.

Two caveats apply. We cannot account for videos removed at upload, nor for ongoing enforcement by TikTok: three accounts were removed by end of collection in April 2026 and a further six were inaccessible by August 2026. We also do not claim that this network is representative of AIGC generally. They are, however, an undeniable part of the TikTok ecosystem: by April 2026, the 4,924 videos had been viewed cumulatively more than 127.9 million times and shared more than 1.28 million times. Some profiles in our dataset retrained followers in the 100,000-1,000,000 range.

Our data highlight the challenging landscape of AIGC labelling. TikTok has acknowledged that off-platform edits and reuploads complicate their detection attempts ^3^, and has announced further work on fraudulent AIGC in the medical space ^4^, which our data suggests is a pressing issue. Notably, comparable challenges appear to face platforms like YouTube, SnapChat, Instagram, Facebook, X and others ^6,7^, but the absence of researcher-friendly data sharing on these sites prevents proper comparison.

## Data Availability

None of the data is available for sharing, in line with requirements of the TikTok Research API Terms of Service.

